# Wastewater-based surveillance for tracing the circulation of Dengue and Chikungunya viruses

**DOI:** 10.1101/2023.10.30.23297765

**Authors:** Sílvia Monteiro, Raquel Pimenta, Filipa Nunes, Mónica V. Cunha, Ricardo Santos

## Abstract

**Background:** Arboviral diseases, transmitted by infected arthropods, pose significant economic and societal threats. Their global distribution and prevalence have increased in recent years, driven by factors such as climate change, biodiversity loss and urbanization. These diseases are often underestimated due to uneven surveillance and asymptomatic cases. Current surveillance relies on monitoring vector occurrence and spatial distribution, as well as syndromic monitoring. In this work, we aimed to explore the utility of wastewater-based surveillance as an additional, added-value tool for vector-borne viruses tracking.

**Methods:** A retrospective wastewater-based surveillance survey was conducted at ten wastewater treatment plants covering a large part of Portugal mainland (North, Lisboa and Vale do Tejo and South regions). Using RT-qPCR, we quantified the RNA concentrations of Dengue (DENV) and Chikungunya (CHIKV) viruses for a period of 12 months (*n* = 273 raw wastewater samples), ranging May 2022 - April 2023.

**Findings:** DENV was detected in 25% of the samples, with concentrations spanning from 3·5 to 6·8 log copies/L. CHIKV was detectable in 11% of the samples, with concentrations up to 6·3 log copies/L. Notably, the occurrence of DENV and CHIKV was rather similar between the three regions. The Lisboa and Vale do Tejo region exhibited in general the highest median concentration for DENV and CHIKV following normalization with crAssphage (1·5 × 10^-4^ and 1·1 × 10^-3^, respectively).

**Interpretation:** We demonstrate the efficacy of wastewater-based surveillance as a potent tool for gauging the epidemiological landscape of both DENV and CHIKV in mainland Portugal, where autochthonous cases have not been detected yet. Therefore, WBS should be integrated as a supplementary component to standard surveillance strategies.

**Research in context:** *Evidence before this study:* Wastewater serves as a valuable resource for wastewater-based surveillance (WBS). This surveillance method involves analyzing biomarkers of human metabolism, activities and lifestyle in wastewater to gain insights into public health trends. Previously, WBS has been employed to track chemical substances like illicit drugs, including cocaine, as well as oseltamivir during the 2009 influenza pandemic. It has also played a crucial role in the global polio eradication program by contributing to assess poliovirus circulation in populations and evaluating immunization effectiveness. During the SARS-CoV-2 pandemic, there was a significant boost in wastewater-based surveillance. To the best of our knowledge, our study represents the first extensive application of this approach to comprehensively investigate and identify the presence of vector-borne viruses RNA such as chikungunya and dengue. In December 2022, we conducted a search on PubMed and Google Scholar for publications using the search terms “wastewater” and “dengue virus” or “chikungunya virus” or “West Nile virus” or “Zika virus”, encompassing manuscripts in all languages. Our search yielded only one study that focused on detecting arboviruses in wastewater, specifically Dengue. Surprisingly, the authors were unable to detect viral RNA despite the reported cases of Dengue between 2017 and 2019, when the wastewater samples were collected. This period included a major outbreak in 2019, with more than 7,000 confirmed cases. Remarkably, no other study has shed light on the presence and concentration of Dengue and Chikungunya viral RNA in wastewater.

*Added value of this study:* In response to the gap identified in the literature review, this study makes a significant contribution by reporting for the first time the presence and concentration of arboviral RNA, specifically focusing on the Dengue and Chikungunya viruses. Notably, it has demonstrated the circulation of these viruses, even when only a few travel-associated cases have been reported throughout the years and where no autochthonous have been reported within the community, although the vector (*Aedes albopictus*) has been identified in several occasions in mainland Portugal. This research underscores the vital role of wastewater-based surveillance in gaining insights into the circulation trends of underreported pathogens within communities at the sewershed level.

*Implications of the available information:* Arboviral infections are significantly underreported, especially in non-endemic countries, and they can lead from mild to severe health issues and even mortality. The symptoms are unspecific and resemble those of various other illnesses, such as Influenza and COVID-19. Consequently, many infected individuals may not seek medical assistance, and when they do, their infections might be misdiagnosed. This results in surveillance for arboviral infections being biased toward individuals presenting with unusual symptoms or severe illness, or returning from endemic regions, leading to delayed reporting and limited temporal and spatial accuracy. In contrast, wastewater surveillance provides results within 24 hours of sample collection and offers a representation of the population served by the sewer system. This includes asymptomatic individuals and those with mild symptoms who may not seek medical care. This approach increases the likelihood of detecting viral circulation and thus prompt public health intervention, namely through vector control, limiting the potential transmission to new hosts, breaking the cycle of transmission. Wastewater surveillance data can play a crucial role in informing public health and environmental decision-making. It can reveal the presence of unaccounted-for infections, indicating the need for control and mitigation strategies, both at the population and mosquito vector levels. Although the arboviral activity is assumed residual in Portugal, wastewater monitoring can be used in complement to the efforts that are already done under the National Vector Surveillance Network. Moreover, this data can be used to communicate with the general public, encouraging them to take actions that can help control the mosquito population, such as reducing stagnant water sources where mosquitoes lay their eggs. This is especially important for vulnerable populations, enabling them to make more informed decisions to mitigate the underlying risks effectively.

## Introduction

Arboviruses, which encompass a diverse range of viruses, including Chikungunya virus (CHIKV) and Dengue virus (DENV), are a heterogeneous group hailing from multiple viral families. These viruses are transmitted to humans through arthropod vectors, primarily mosquitoes and ticks. The array of arboviruses linked to human diseases surpasses a hundred, resulting in considerable disease burden and socioeconomic implications, especially in tropical and subtropical regions^1^. Presently, arboviruses pose a global public health concern due to the influences of climate change, globalization, biodiversity loss, landscape homogenization and urbanization^2,3^.

Among arboviruses, DENV stands out as the most clinically significant, as it is endemic in over 100 countries^4^. Its far-reaching presence contributes to an estimated 390 million infections annually, with only approximately 25% of these cases reporting symptoms^5^. Conversely, CHIKV has instigated extensive outbreaks across Africa, Asia, and the Americas; however, it has subsequently extended its reach to regions where previously it was not native, including Europe^6^.

Assessing the presence of arboviruses such as DENV and CHIKV involves multiple approaches, including clinical surveillance and monitoring within the sylvatic and entomological cycles. Clinical surveillance remains the primary method for tracking arboviral diseases. However, it is estimated that over 50% of arboviral infections may be asymptomatic or exhibit mild symptoms^7^. Additionally, non-severe arboviral infections often manifest with nonspecific symptoms, complicating early outbreak detection^8^.

On the other hand, monitoring arbovirus circulation in the sylvatic and entomological cycles is also commended. However, this approach presents challenges, such as the need to capture animals and trap insect reservoirs. Moreover, using the presence of insect reservoirs as proxy for human infections may not reflect the epidemiological scenario^9^. Therefore, both clinical surveillance and vector cycle-based monitoring lack sensitivity and specificity, are resource-intensive, and lack spatial precision^10,11^.

The Asian tiger mosquitoes, *Aedes albopictus* and *Aedes aegypti*, act as the primary vectors of DENV and CHKV. *A. albopictus* was detected for the first time in mainland Portugal, in the North region, by the end of July 2017, and associated to international trade of used tires that can be used by mosquito as oviposition site. Detection in the Algarve region was subsequently reported. Imported dengue cases have been identified in mainland Portugal, although there is no evidence of local transmission (SNS 24: https://www.sns24.gov.pt/tema/doencas-infecciosas/dengue/#vivo-em-portugal-devo-preocupar-me-com-o-risco-de-infecao-pelo-virus-da-dengue. Last accessed: 30 august 2023).

Given the limitations of current surveillance methodologies for arboviral diseases concerning particularly the asymptomatic and mild cases, there is a pressing need for rapid, near-real-time approaches to determine the extent of arboviral circulation in human populations. Wastewater-based surveillance (WBS) has successfully been used to trace pharmaceuticals, illicit drugs, and even polio. It was extensively employed during the severe acute respiratory syndrome coronavirus 2 (SARS-CoV-2) pandemic ^13^. Studies have confirmed the excretion of DENV and CHIKV RNA in urine at concentrations as high as 5·2 log_10_ copies/mL for DENV.^14^ Additionally, research has indicated that DENV RNA can persist for several days at temperatures as high as 37 ºC, suggesting that monitoring of viral RNA in wastewater could complement clinical surveillance for arboviral diseases^15^.

In this study, we present a nation-wide analysis of DENV and CHIKV RNA in raw wastewater across Portugal over a 54-week period. The CrAssphage served as a normalization factor to adjust the arboviral concentrations, taking into consideration the varying dilution rates of raw urban wastewater and the shifting proportion of human fecal material over time.^16^ These factors, influenced by stormwater, industrial discharges, and water infiltration, could potentially lead to misinterpretations when utilizing WBS. The insights gained from this research will advance our understanding of the potential underlying wastewater-based surveillance for monitoring arboviral outbreaks, including those caused by DENV and CHIKV, especially in non-endemic regions.

## Methods

### Study design

Twice a month, raw wastewater samples (n = 273) were systematically collected from ten wastewater treatment facilities situated across the North (*n =* 75 samples), Lisboa and Vale do Tejo (*n* = 98), and South (*n* = 100) regions of Portugal, from May 2022 to April 2023 (appendix page 7). Employing automated samplers (ISCO, USA), 24-hour composite samples were collected. Following collection, the samples were promptly refrigerated and transported to the laboratory within an 8-hour window, where they were immediately processed.

### Processing of raw wastewater

To concentrate the samples, a hollow fiber method following the procedure outlined by Monteiro and colleagues was employed (appendix pp 2) ^17^. In brief, 1 liter of raw wastewater underwent concentration using Inuvai R180 hollow-fiber filters (Inuvai, a division of Fresenius Medical Care, Germany). The elution of samples was conducted in 300 mL of 1x phosphate-buffered saline (PBS) supplemented with 0·01% sodium polyphosphate (NaPP) and 0·01% Tween 80/0·001% antifoam, with subsequent overnight precipitation with 20% polyethylene glycol (PEG) 8000. Samples were subjected to centrifugation at 10,000 xg for 30 minutes, followed by resuspension in 5 mL of 1x PBS at pH 7·4. The samples were stored at (−80 ± 10) °C until further processing.

### Viral RNA extraction, detection, and quantification

Nucleic acid extraction was carried out on 220 μL of concentrated samples using the QIAamp FAST DNA Stool Mini kit (QIAGEN, Germany) with a final elution volume of 100 μL, following the manufacturer’s instructions (appendix pp 2).

The primers and probes employed in this study can be found in Table 1. For DENV RNA detection, the focus was on a segment in the 3’-untranslated region (3’UTR), while CHIKV RNA detection targeted both the Nsp1 and the E region^18-20^. Quantification of crAssphage was executed using the previously established CPQ_056 assay^21^.

**Table 1:**
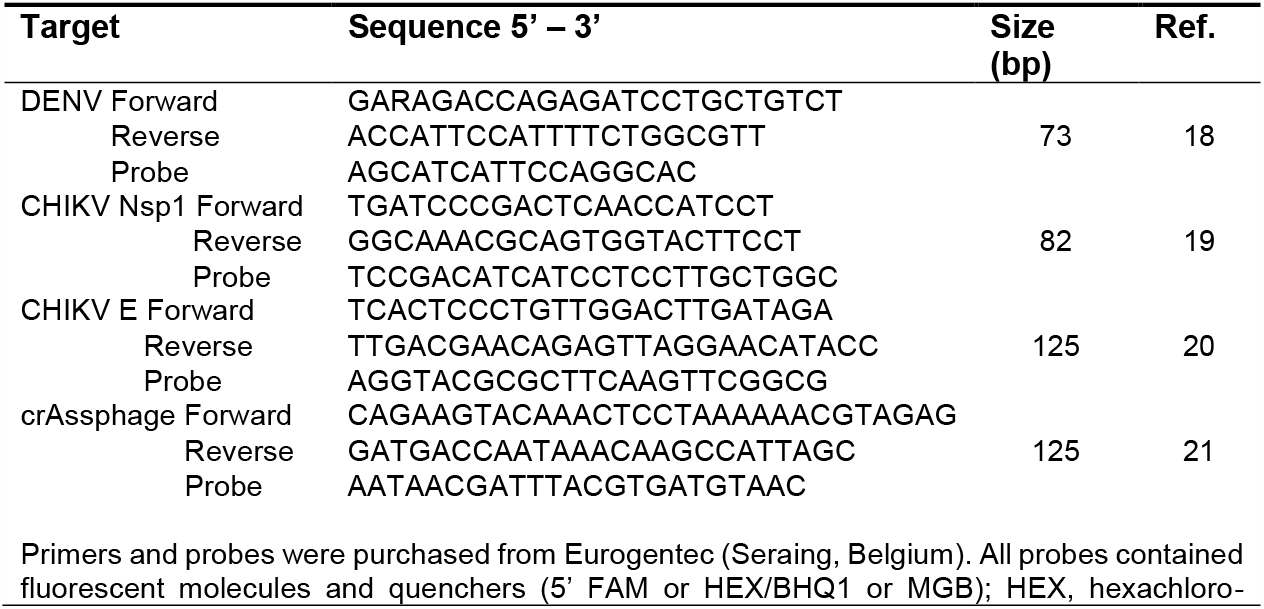
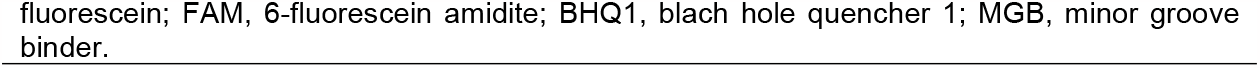
Forward and reverse primers and probe sequences used in this study for the detection of viral RNA/DNA.

Detection of DENV and CHIKV involved one-step RT-qPCR assays (Luna Universal Probe One-Step RT-qPCR, New England Biolabs, USA), while qPCR assay (PerfeCTa qPCR ToughMix, Quantabio, USA) was used for crAssphage quantification. To detect viral nucleic acids, 4-fold dilutions of each viral extract were concurrently assayed alongside crude extracts to overcome any potential inhibition of amplification caused by the complex nature of the samples.

The reaction mixture’s final volume was 15 μL, consisting of 800 nM of each primer, 200 nM of probe, and 5 μL of the extracted viral nucleic acid. RT-qPCR for DENV and CHIKV followed a temperature profile of 55 ºC for 10 min, 95 ºC for 10 min, and 45 cycles of amplification at 95 ºC for 15 s, and 60 ºC for 1 min. The qPCR for crAssphage was conducted at 95 ºC for 10 min, followed by 45 cycles of amplification at 95 ºC for 15 s, and 60 ºC for 1 min. All amplification reactions were performed using a QuantStudio 5 Real-Time PCR System (ThermoScientifc, USA).

Quantification of DENV and CHIKV was achieved through calibration curves utilizing 5- and 10-fold dilutions of the TaqMan Arbovirus Triplex Control Kit (ZIKV/DNV/CHIKV) (ThermoScientific, USA), ranging from 6·0 to 6·0×10^3^ and 1·0 to 1·0×10^4^ genome copies (gc) per reaction. Standard curves for crAssphage were generated from genomic crAssphage DNA and quantified as per the guidelines in appendix (pp 2). The performance details of the standard curves can be found in appendix pp 3. Negative controls, including extraction and RT-qPCR assays, were also carried out using DNase/RNase-free distilled water (appendix pp 4-6).

### Validation of arboviral detection in wastewater by sequencing

To validate the detection of DENV and CHIKV in wastewater samples, the amplicons were visualized after electrophoresis on a 4·0% agarose gel, and ethidium staining. The bands with the correct size (Table 1) were excised from the gel and extracted using the EXTRACTME DNA clean-up gel-out kit (Blirt, Poland) following the manufacturer’s instructions. The DNA was recovered in final volume of 50 μL. Positive controls for both viruses were also sequenced in parallel with the samples. The sequencing was conducted by an external laboratory. Upon receiving the sequences, the results were checked and compared with publicly available sequences by using the BLAST program of the National Center for Biotechnology Information (available from: URL: https://blast.ncbi.nlm.nih.gov/Blast.cgi?PROGRAM=blastn&BLAST_SPEC=GeoBlast&PAGE_TYPE=BlastSearch).

### Statistical analysis

Data analysis was conducted using SPSS version 26 (IBM Corporation, USA) and RStudio (version 2023.06.1-524). Figures were generated with RStudio, employing the ggplot2 package (version 3.4.0).

In preparation for statistical analyses, all positive data underwent a log transformation. Quantitative analysis was exclusively performed on positive samples, with non-detects (ND) being excluded from quantitative calculations. The normality of the data was assessed using the Shapiro-Wilk test. The study employed the Kruskal-Wallis test (KW statistics) to assess variations in viral RNA concentration across different regions and seasons. Additionally, Spearman’s rank correlation coefficient was utilized to identify potential relationships between the three viral targets (DENV, CHIKV – Nsp1, and CHIKV – E). Differences were considered statistically significant if *p* < 0·05.

## Results

The control experiments yielded the anticipated outcomes (including negative and positive extraction and amplification controls). The median concentration of crAssphage, utilized for normalizing arboviral concentration across all samples, was 6·8 × 10^8^ copies/L, with a range of 4·4 × 10^6^ to 5·5 × 10^9^ copies/L and an interquartile range between 2·7 × 10^8^ and 1·2 × 10^9^ copies/L. Our study’s crAssphage concentration results align with findings from previously published studies.^22,23^

The RNA from all viruses under scrutiny was consistently detected in raw urban wastewater samples (see Figure 1). DENV was consistently present throughout the sampling period, while CHIKV detection was more prevalent in 2023, particularly during March and April. Specifically, DENV was found in 68 samples (25%), while CHIKV was detected in 30 samples (11%). Among the samples positive for arboviral RNA, 62 (67%) were positive for DENV, 24 (26%) for CHIKV, and, overall, both viruses were concomitantly detected in six (7%) of raw urban wastewater samples.

**Figure 1:**
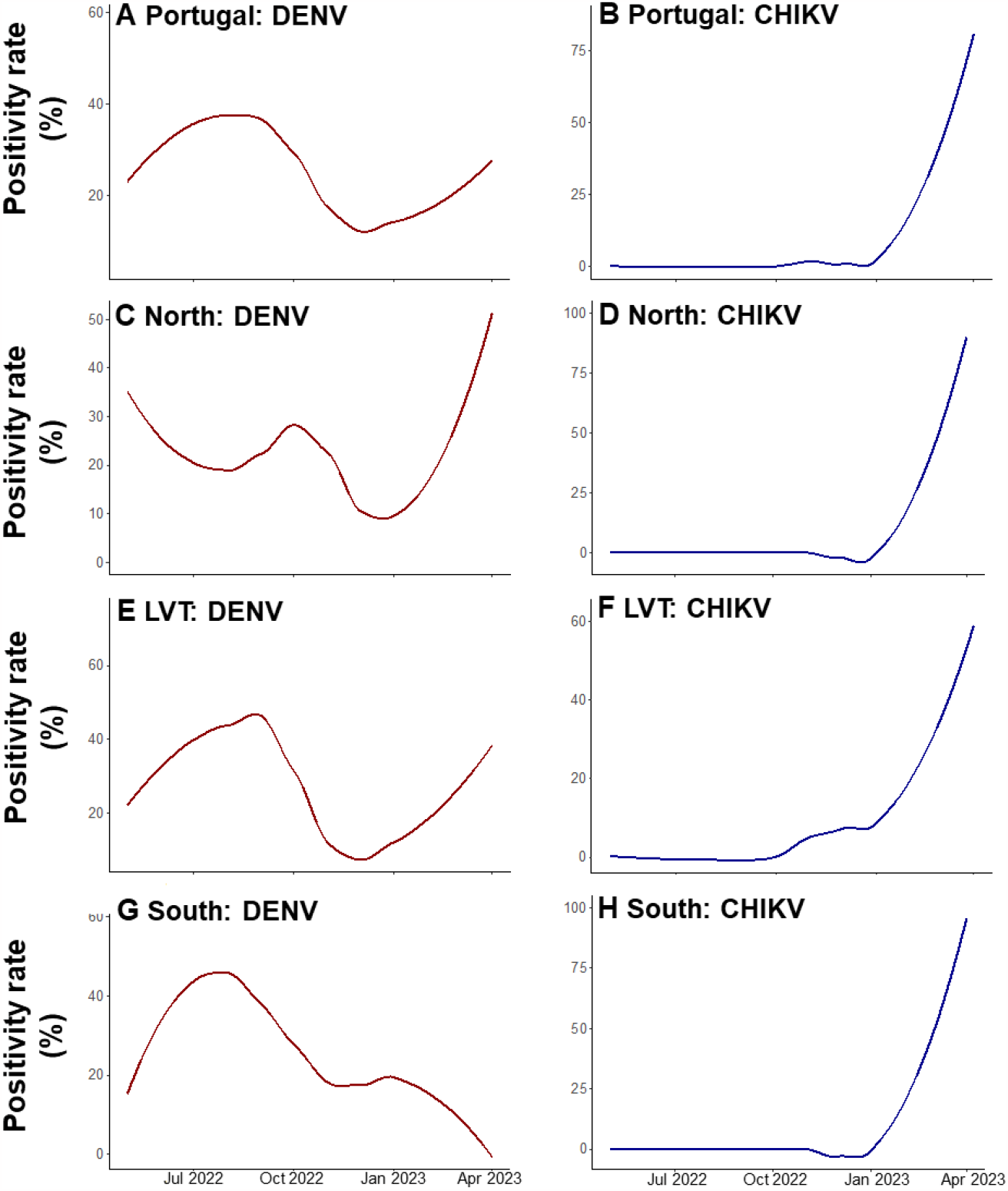
Portugal- and Portuguese regions-aggregated positivity rates in raw urban wastewater. Report of bimonthly measurements. (A) Positive rate for DENV in Portugal. (B) Positive rate for CHIKV in Portugal, includes the genes Nsp1 and E. (C) Positivity rate for DENV in the North region; (D) Positivity rate for CHIKV in the North region (Nsp1 and E). (E) Positivity rate for DENV in the LVT region; (F) Positivity rate for CHIKV in the LVT region (Nsp1 and E). (G) Positivity rate for DENV in the South region. (H) Positivity rate for CHIKV in the South region (Nsp1 and E). Measurements were conducted between May 2022 and April 2023. DENV = dengue virus. CHIKV = chikungunya virus.

Regarding DENV, the highest positivity rate during the study period occurred in September 2022 (59%), and the lowest was in January 2023 (4%). As for CHIKV, the first detection was observed in a single sample from the LVT region in December 2022 (see Figure 1F), and the highest positivity rate was noted in April 2023. The trends for DENV varied slightly across the three regions. Initially, the highest detection rate occurred in the North and LVT regions (33%). The positivity rate in the North region decreased until August, while an increase in positive samples was registered for the LVT and South regions until September 2022, with rates reaching up to 75% for LVT and 63% for the South region. Two additional peaks were observed in the North region, in September and October 2022, as well as between February and April 2023. In the LVT region, there was a sharp decrease in positivity rate after October 2022, followed by an increase from February 2023 onward. In the South region, a peak was detected between December 2022 and February 2023, with no positive samples in March and April 2023.

For CHIKV, the LVT region showed the first positive samples in December 2022, with subsequent detections in January (13%), March (38%), and April (63%). The North and South regions had their first positive samples in March 2023 (33% and 50%, respectively), with all April samples testing positive in both regions.

The normalized concentrations ranged between a median of 1·1 × 10^−4^ and 7·8 × 10^−4^ copies/L for DENV and the CHIKV gene E (see Figure 2; Table 2).

**Table 2:**
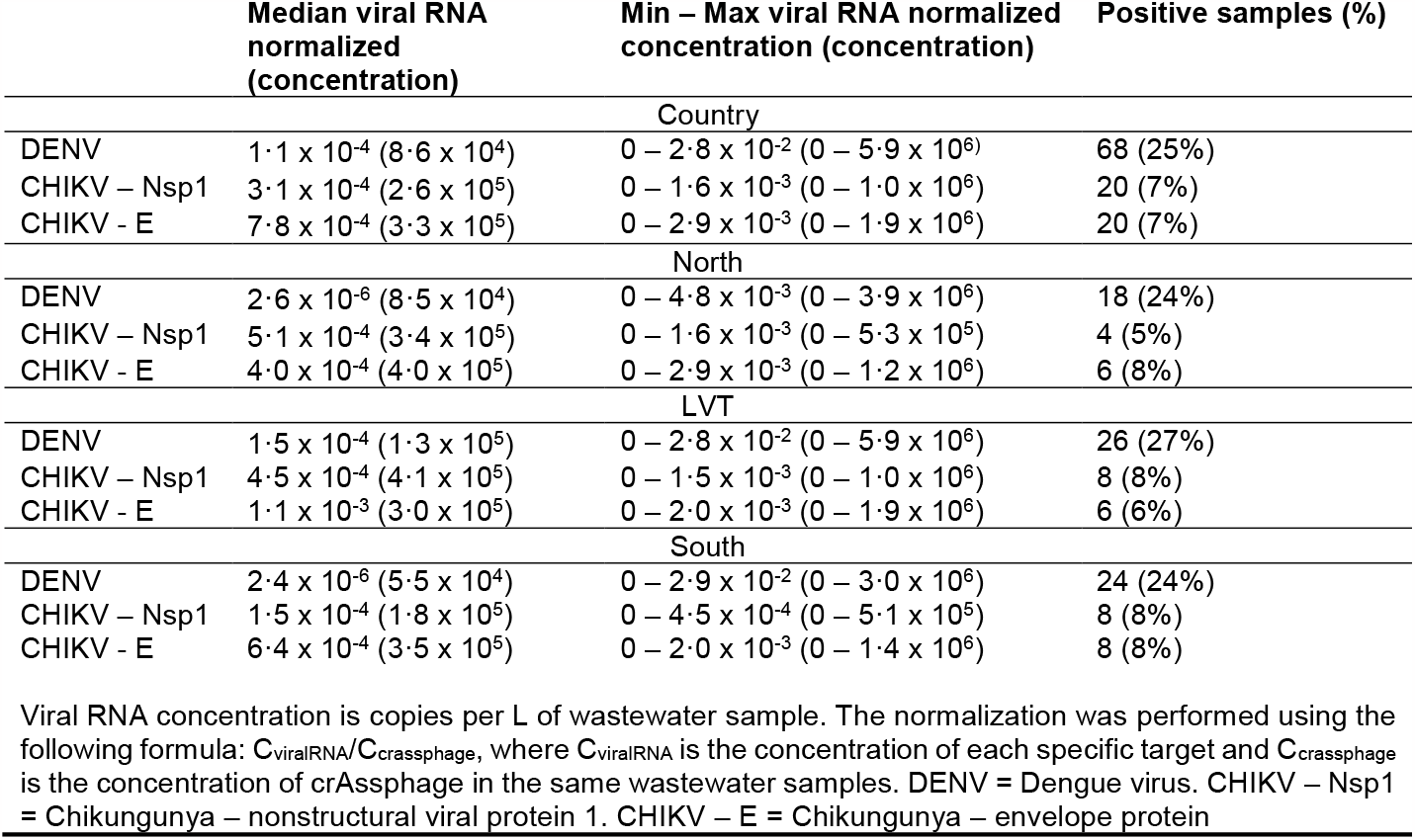
Summary statistics of the detection of DENV and CHIKV in wastewater samples

|  | Median viral RNA normalized (concentration) | Min – Max viral RNA normalized concentration (concentration) | Positive samples (%) |
| --- | --- | --- | --- |
| Country |  |  |  |
| DENV | $1.1 \times 10^{-4}$ ($8.6 \times 10^4$) | $0 - 2.8 \times 10^{-2}$ ($0 - 5.9 \times 10^6$) | 68 (25%) |
| CHIKV – Nsp1 | $3.1 \times 10^{-4}$ ($2.6 \times 10^5$) | $0 - 1.6 \times 10^{-3}$ ($0 - 1.0 \times 10^6$) | 20 (7%) |
| CHIKV - E | $7.8 \times 10^{-4}$ ($3.3 \times 10^5$) | $0 - 2.9 \times 10^{-3}$ ($0 - 1.9 \times 10^6$) | 20 (7%) |
| North |  |  |  |
| DENV | $2.6 \times 10^{-6}$ ($8.5 \times 10^4$) | $0 - 4.8 \times 10^{-3}$ ($0 - 3.9 \times 10^6$) | 18 (24%) |
| CHIKV – Nsp1 | $5.1 \times 10^{-4}$ ($3.4 \times 10^5$) | $0 - 1.6 \times 10^{-3}$ ($0 - 5.3 \times 10^5$) | 4 (5%) |
| CHIKV - E | $4.0 \times 10^{-4}$ ($4.0 \times 10^5$) | $0 - 2.9 \times 10^{-3}$ ($0 - 1.2 \times 10^6$) | 6 (8%) |
| LVT |  |  |  |
| DENV | $1.5 \times 10^{-4}$ ($1.3 \times 10^5$) | $0 - 2.8 \times 10^{-2}$ ($0 - 5.9 \times 10^6$) | 26 (27%) |
| CHIKV – Nsp1 | $4.5 \times 10^{-4}$ ($4.1 \times 10^5$) | $0 - 1.5 \times 10^{-3}$ ($0 - 1.0 \times 10^6$) | 8 (8%) |
| CHIKV - E | $1.1 \times 10^{-3}$ ($3.0 \times 10^5$) | $0 - 2.0 \times 10^{-3}$ ($0 - 1.9 \times 10^6$) | 6 (6%) |
| South |  |  |  |
| DENV | $2.4 \times 10^{-6}$ ($5.5 \times 10^4$) | $0 - 2.9 \times 10^{-2}$ ($0 - 3.0 \times 10^6$) | 24 (24%) |
| CHIKV – Nsp1 | $1.5 \times 10^{-4}$ ($1.8 \times 10^5$) | $0 - 4.5 \times 10^{-4}$ ($0 - 5.1 \times 10^5$) | 8 (8%) |
| CHIKV - E | $6.4 \times 10^{-4}$ ($3.5 \times 10^5$) | $0 - 2.0 \times 10^{-3}$ ($0 - 1.4 \times 10^6$) | 8 (8%) |

Viral RNA concentration is copies per L of wastewater sample. The normalization was performed using the following formula: $C_{\text{viralRNA}}/C_{\text{crassphage}}$ , where $C_{\text{viralRNA}}$ is the concentration of each specific target and $C_{\text{crassphage}}$ is the concentration of crAssphage in the same wastewater samples. DENV = Dengue virus. CHIKV – Nsp1 = Chikungunya – nonstructural viral protein 1. CHIKV – E = Chikungunya – envelope protein

**Figure 2:**
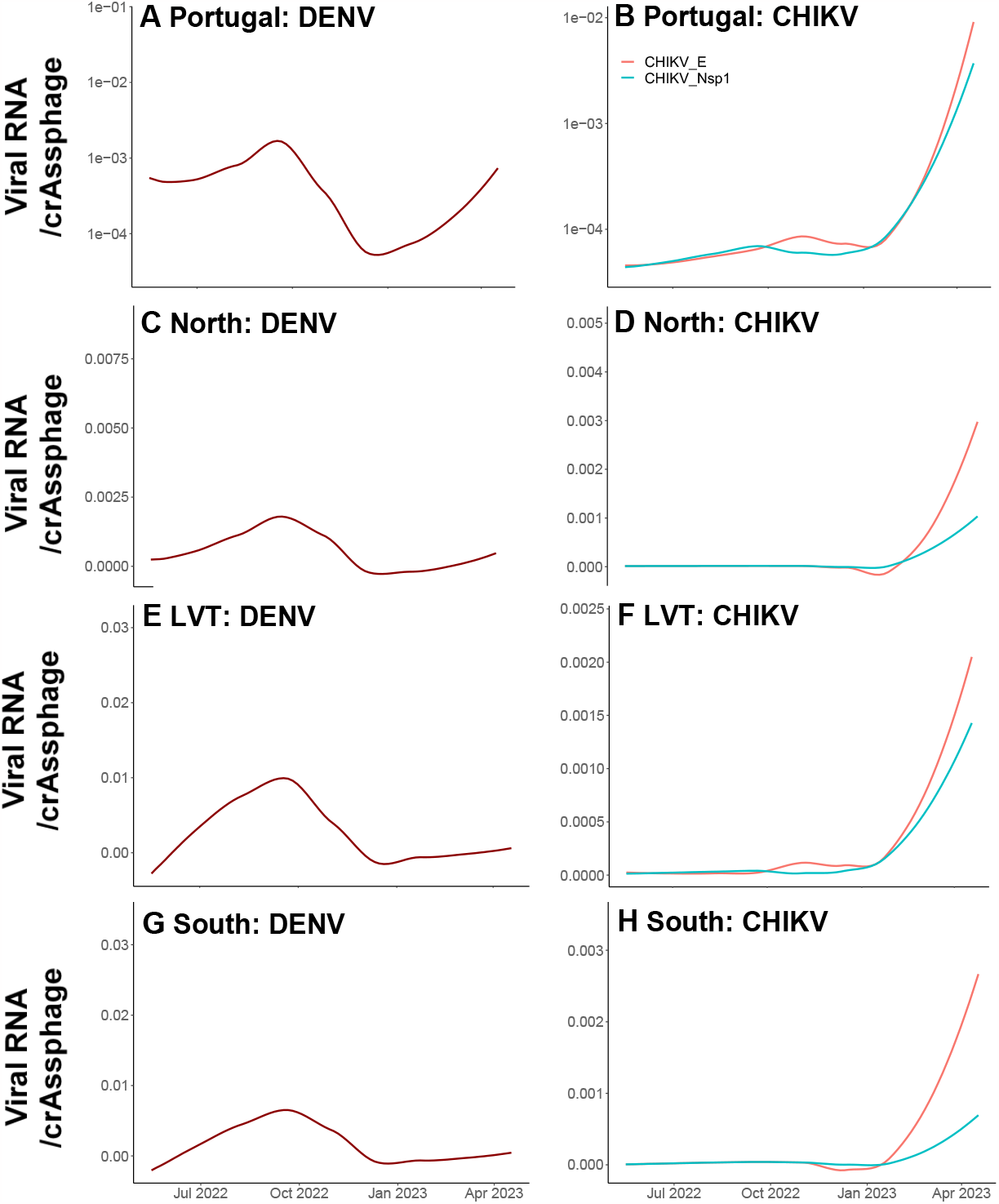
Portugal- and Portuguese regions-aggregated normalized concentration of arboviral RNA in urban raw wastewater Report of bimonthly measurements. (A) Normalized concentration of DENV in Portugal. (B) Normalized concentration of CHIKV in Portugal, expressed by the genes Nsp1 and E. (C) Normalized concentration of DENV in the North region; (D) Normalized concentration of CHIKV in the North region (expressed by the Nsp1 and E genes). (E) Normalized concentration of DENV in the LVT region; (F) Normalized concentration of CHIKV in the LVT region (expressed by the Nsp1 and E genes). (G) Normalized concentration of DENV in the South region. (H) Normalized concentration of CHIKV in the South region (expressed by the Nsp1 and E genes). Measurements were conducted between May 2022 and April 2023. DENV = dengue virus. CHIKV = chikungunya virus.

The median normalized concentration of the CHIKV Nsp1 gene was similar (3·1 × 10^−4^ copies/L). Generally, the South region exhibited the lowest average concentrations, while the LVT region had the highest. For DENV, the normalized concentration varied between 2·4 × 10^−6^ copies/L in the South region and 1·5 × 10^−4^ copies/L in the LVT region. The concentration of the E gene followed a similar trend, with the highest median concentration (1·1 × 10^−3^ copies/L) in the LVT region and the lowest (4·0 × 10^−4^ copies/L) in the North region. However, the concentration for the Nsp1 gene was very similar across regions (ranging from 1·5 × 10^−4^ to 5·1 × 10^−4^ copies/L for the South and North regions, respectively). No statistically significant differences were observed in the normalized RNA viral concentration among the three locations (DENV: KW = 2·738, degrees of freedom = 2, p = 0·25; CHIKV – Nsp1: KW = 3·864, degrees of freedom = 2, p = 0·15; CHIKV – E: KW = 0·437, degrees of freedom = 2, p = 0·80).

In general, the concentration of DENV was lowest between the end of October 2022 and the beginning of January 2023, with the highest concentrations observed between September and October 2022, and a resurgence at the start of 2023, except for the South region where DENV remained mostly undetected. For CHIKV, the virus was primarily undetected during 2022 but saw a steep increase from January until the end of the time series. The non-normalized concentration followed a similar trend to that of the normalized concentration for both viruses (appendix pp 8). Concentrations of both CHIKV genes were positively correlated with each other (r = 0·44, p < 0·001), while no significant association was found between DENV and CHIKV genes (appendix pp 9). The limited occurrence of CHIKV throughout the sampling period contributed to this result.

The impact of the four different seasons on the occurrence of viral RNA was evaluated. Statistically significant differences were observed for all targets (DENV, CHIKV – Nsp1, and CHIKV – E) (p < 0·001). For DENV, the difference was notable between warmer seasons (spring and summer) and colder seasons (fall and winter), with higher concentrations and positivity rates found during the warmer months (appendix pp 9, 10). Conversely, CHIKV was more dominant during spring, with a statistically significant difference observed between spring and the other seasons (appendix pp 9, 10).

The sequencing data, although not being able to provide discrimination between viruses (in the case of dengue, serotypes) and therefore not being able to establish a phylogenetic tree, due to the small size of the fragments, confirmed the detection of each virus in the positive samples.

## Discussion

To the best of our knowledge, this is the first study to document the presence of RNA from DENV and CHIKV in raw urban wastewater samples.

DENV was detected throughout the study whereas CHIKV RNA was seldomly detected in wastewater. CHIKV and DENV co-occurred in six samples (7%). Both viruses co-circulate in many regions of the globe, implying that these viruses can be spread simultaneously to other regions where the viruses are not endemic (ECDC: https://www.ecdc.europa.eu/en/mosquito-borne-diseases. Last accessed: October 2023). Moreover, the co-circulation of both viruses and of the vectors facilitates the occurrence of concurrent infection in the human population.^24^ An association between viral RNA in raw urban wastewater and clinical surveillance could not be established, given the absence of data, encompassing both autochthonous and travel-associated cases, of CHIKV and DENV in Portugal from August 2022 to July 2023 (Turismo de Portugal: https://travelbi.turismodeportugal.pt/turismo-em-portugal/turismo-em-numeros-2022/. Last accessed: October 2023).

Between 2015 and 2021, Portugal reported only three cases of CHIKV and 96 cases of DENV, all travel-associated (ECDC: https://www.ecdc.europa.eu/en/surveillance-atlas-infectious-diseases. Last accessed: October 2023). However, changing climate, which favors vector activity and viral replication, coupled with increased globalization (including travel, migration, and international trade), is expected to lead to a rise in CHIKV and DENV cases across Europe. In fact, the *A. albopictus* is presently expanding across Europe temperate and Mediterranean regions, being established in Italy, France, and Spain, among other countries, where it has mostly been associated to intermittent and short-lived outbreaks rather than continuous transmission (ECDC: https://www.ecdc.europa.eu/en/publications-data/aedes-albopictus-current-known-distribution-august-2023. Last accessed: October 2023).

DENV exhibited its highest concentration and positivity rate in September 2022, with a resurgence between March and April 2023. In contrast, CHIKV was predominantly detected from January 2023 onward. There is no discernible seasonality associated with either virus, and the peak in the number of cases in Europe varies from year to year, influenced by climatic conditions and fluctuating travel patterns (The European Climate Adaptation Platform Climate-ADAPT: https://climate-adapt.eea.europa.eu/. Last accessed: October 2023). This lack of seasonality pattern may be due to the high ecological adaptability and plasticity of *A. albopictus*, with cold-acclimated adult mosquitoes and drought-resistant eggs.^25^

Portugal, being a significant tourist destination, experienced a substantial influx of international guests, with peaks in July, August, September, and October. This surge in tourism, equivalent to approximately 20% of the country’s population in these months, may have a notable impact on the presence of DENV and CHIKV (Turismo de Portugal: https://travelbi.turismodeportugal.pt/turismo-em-portugal/turismo-em-numeros-2022/. Last accessed: October 2023). Portugal also attracted an increasing number of immigrants, including digital nomads from countries such as Brazil, the USA, and the UK, potentially contributing to the circulation of these viruses. Moreover, Portugal has direct flight around the year from Sub-Saharan Africa and Latin America, specially from countries that are former Portuguese territory.

In addition to its status as a significant tourism hub, Portugal has also emerged as an enticing destination for permanent residence (Instituto Nacional de Estatística: https://www.ine.pt/xportal/xmain?xpid=INE&xpgid=ine_publicacoes&PUBLICACOESpub_boui=65586079&PUBLICACOESmodo=2. Last accessed: October 2023). Between December 2021 and December 2022, Portugal’s population experienced a notable increase of 46,249 individuals, primarily driven by a net migration influx of nearly 87,000 people. This marked an uptick of nearly 15,000 migrants compared to the previous year (DGS: https://www.dgs.pt/em-destaque/mosquito-aedes-albopictus-identificado-no-municipio-de-lisboa.aspx. Last accessed: October 2023).

Notably, the SARS-CoV-2 pandemic shifted established work norms towards remote employment, leading to a surge in digital nomads choosing Portugal as their touch base. Census data for the year 2021 reveals that Brazil, Italy, and India ranked among the top five nationalities of immigrants, with Brazil comprising nearly 30% of the total immigrant population (Instituto Nacional de Estatística: https://www.ine.pt/xportal/xmain?xpid=INE&xpgid=ine_publicacoes&PUBLICACOESpub_boui=65586079&PUBLICACOESmodo=2. Last accessed: October 2023). Additionally, the number of immigrants from South Asian nations like India and Pakistan has experienced a steep rise in the mainland territory for the past two years since the last census. Major Portuguese cities, including Lisbon, Porto, Setúbal, Faro, and Braga, situated within the regions under study, attract the majority of immigrants and also host the highest influx of tourists. Consequently, the interplay between tourism to and from Portugal, coupled with the escalating immigrant population, may have played a role in influencing the positivity rates and concentrations of both viruses in the region.

Before this study, in the scope of another work particularly focusing on the fate of SARS-CoV-2 across the water cycle, we have detected, via shotgun viral metagenomic sequencing by Illumina, a densovirus (*Brevidensovirus* genus, subfamily *Densovirinae*) specific of *A. albopictus* (Cunha *et al*., unpublished data). Detection of this densovirus in raw wastewater from the North, Algarve and LVT regions of Portugal was confirmed for a number of samples collected during 2021. Mosquito densoviruses exert mosquito-specific entomopathogenicity and their detection may also be used as proxy of mosquito occurrence. According to the ECDC, the distribution of *A. albopictus* in the mainland territory of Portugal between March 2021 and August 2023 was confined to the North (Penafiel) and the Algarve regions (ECDC: https://www.ecdc.europa.eu/en/publications-data/aedes-albopictus-current-known-distribution-august-2023. Last accessed: October 2023). However, using metagenomic detection of *A. albopictus* densivirus in wastewater as proxy for the presence of the mosquito vector, wastewater analyses was able to extend information on the spatial distribution of *A. albopictus* across the mainland territory, confirming for the first time its occurrence in the Lisbon area already back in 2021, a fact that only in 2023 was confirmed via classical mosquito trapping methods (DGS: https://www.dgs.pt/em-destaque/mosquito-aedes-albopictus-identificado-no-municipio-de-lisboa.aspx. Last accessed: October 2023). This additional data supports the utility of WBS for understanding the circulation trends of arboviral pathogens and their vectors.

Despite the absence of reported DENV and CHIKV cases in Portugal, uncertainties persist regarding disease occurrence when testing wastewater. Information on the concentration of viral particles excreted by infected individuals, whether symptomatic or asymptomatic, or the concentration of these viruses in the vectors is limited and challenging to obtain. Wastewater surveillance is not suitable for intervention at the individual level and cannot discriminate infection patterns from permanent and transient populations.

Nonetheless, this study successfully identified the presence and concentration of DENV and CHIKV, even in the absence of data for reported cases. The lack of reporting can be attributed to several factors, including the absence of symptoms, the presence of mild symptoms, or misdiagnosis. For example, it is estimated that only around 25% of DENV cases exhibit symptoms.^5^ Moreover, both DENV and CHIKV can cause mild and common symptoms, such as fever, headache, muscle pain, and fatigue, which are also typical of SARS-CoV-2, influenza or other entero- or neurotropic viruses. This leads to a situation where asymptomatic individuals may not seek medical care, and even when they do, they may receive a misdiagnosis.

Continuous and active surveillance remains indispensable to timely detect and manage potential outbreaks caused by Dengue and Chikungunya viruses. The detection of *A. albopictus* densiviruses across WWTP via unbiased metagenomic sequencing, together with tailored molecular diagnostic tests targeting the genome of specific arboviruses in wastewater, enrich the available surveillance toolbox for relevant arboviral diseases, which traditionally has been circumscribed to syndromic surveillance and vector trapping/testing. These novel approaches combining the information pool from wastewater and clinical samples amplify methodological choices for predicting the infection risk, aiding to prevent and mitigate potential Dengue and Chikungunya outbreaks. Furthermore, since mosquito surveillance and control remain the most effective strategy to prevent and reduce arboviruses transmission, integrating vector occurrence and distribution from wastewater data with ongoing harmonized surveillance of *Aedes* invasive mosquito species is crucial to mitigate human vector-borne diseases.

Further research is warranted to comprehensively understand the levels of arboviral excretion by symptomatic and asymptomatic individuals, patterns of infection, and vector dynamics. Such knowledge would enable accurate modeling of the number of infected individuals within a sewer system based on viral concentration, thereby facilitating the modeling of epidemiological parameters.^26-27^ Nevertheless, WBS is poised to become an even more critical tool in combating arboviral diseases, both in endemic and non-endemic regions, as these areas are expected to become increasingly susceptible to diseases such as dengue and chikungunya.

## Data Availability

All data produced in the present study are available upon reasonable request to the authors

## Contributors

SM contributed to the conceptualization of the study, the resources, data curation, writing of the original draft, reviewing and editing of subsequent drafts, visualization, and supervision. RP and FN contributed to the investigation. MVC contributed to reviewing and editing of the manuscript drafts. RS contributed to the conceptualization of the study, the resources, data curation, writing of the original draft, reviewing and editing of subsequent drafts, visualization, and supervision. All authors had access to all the data in the study and had final responsibility for the decision to submit for publication. SM and RS verified the underlaying data of the study.

## Acknowledgments

We acknowledge the institutional support of *Ministério do Ambiente e Ação Climática* and of the *Direção Geral de Saúde*. We thank all the operational workers from water utilities [Águas de Portugal Group, AGERE and SMAS de Almada] who contributed to wastewater sampling.

## References

1 LaBeaud AD, Bashir F, King CH. Measuring the burden of arboviral diseases: the spectrum of morbidity and mortality from four prevalent infections. Popul Health Metr 2011; 9: 1. doi: 10.1186/1478-7954-9-1

2 Garnas J, Auger-Rozenberg MA, Roques A, et al. Complex patterns of global spread in invasive insects: eco-evolutionary and management consequences. Biol Invasions 2016; 18: 935–952. Doi: 10.1007/s10530-016-1082-9

3 Robert MA, Stewart-Ibarra AM, Estallo E. Climate change and viral emergence from Aedes-borne arboviruses. Curr Opin Virol 2020; 40: 41–47.

4 Messina JP, Brady OJ, Scott TW, et al. Global spread of Dengue virus types: mapping the 70 year history. Trends Microbiol 2014; 22: 138–146. Doi: 10.1016/j.tim.2013.12.011

5 Bhatt S, Gething PW, Brady, OJ, et al. The global distribution and burden of dengue. Nature 2013; 496: 504–507. Doi: 10.1038/nature12060

6 Guzman MG, Gubler DJ, Izquierdo A, Martinez E, Halstead SB. Dengue infection. Nat Rev Dis Prim 2016; 2: 16055. Doi: 10.1038/nrdp.2016.55

7 Wahid B, Ali A, Rafique S, Idrees M. Global expansion of chikungunya virus: mapping the 64-year history. Int J Infect Dis 2017; 58: 69–76. Doi: 10.1016/j.ijid.2017.03.006

8 Yew YW, Ye, T, Ang LW, et al. Seroepidemiology of Dengue virus infection among adults in Singapore. Ann Acad Med Singap 2009; 38: 667–675

9 Musso D, Rodriguez-Morales AJ, Levi JE, Cao-Lormeau VM, Gubler DJ. Unexpected outbreaks of arbovírus infections: lessons learned from the pacific and tropical America. Lancet Infect Dis 2018; 18: e355–e361. doi: 10.1016/S1473-3099(18)30269-X

10 Bowman LR, Runge-Ranzinger S, McCall PJ. Assessing the relationship between vector indices and dengue transmission: a systematic review of the evidence. PLoS Negl Trop Dis 2014, 8: e2848. Doi: 10.1371/journal.pntd.0002848

11 Tambo E, Khayeka-Wandabwa C, Olalubi AO, Adedeji AA, Ngogang JY, Khater EIM. Addressing knowledge gaps in molecular, sero-surveillance and monitoring approaches on Zika epidemics and other arbovirus co-infections: a structured review. Parasite Epidemiol Control 2017; 2: 50–60. doi: 10.1016/j.parepi.2017.01.001

12 European Centre for Disease Prevention and Control. Dengue outbreak in Madeira, Portugal, March 2013. Stockholm, ECDC, 2014.

13 Naughton C. COVIDPoops19 Summary of Global SARS-CoV-2 wastewater monitoring efforts. https://ucmerced.maps.arcgis.com/apps/dashboards/c778145ea5bb4daeb58d31afee389082 last accessed: 30 august 2023)

14 Andries AC, Duong V, Ly S, et al. Value of routine dengue diagnostic tests in urine and saliva specimens. PLos Negl Trop Dis 2015; 9: e0004100. doi: 10.1371/journal.pntd.0004100

15 Chandra F, Lee WL, Armas F, et al. Persistence of Dengue (serotypes 2 and 3), Zika, Yellow Fever, and murine hepatitis virus RNA in untreated wastewater. Environ Sci Technol Lett 2021; 8: 785–791. Doi: 10.1021/acs.estlett.1c00517

16 Wilder ML, Middleton F, Larsen DA, et al. Co-quantification of crAssphage increases confidence in wastewater-based epidemiology for SARS-CoV-2 in low prevalence areas. Water Res. X 2021; 11: 100100. doi: 10.1016/j.wroa.2021.100100

17 Monteiro S, Rente D, Cunha MC, et al. A wastewater-based epidemiology tool for COVID-19 surveillance in Portugal. Sci Total Environ 2022; 804: 150264. doi: 10.1016/j.scitotenv.2021.150264

18 Gurukumar KR, Priyadarshini D, Patil JA, et al. Development of a real time PCR for detection and quantification of Dengue viruses. Virol J 2009; 6: 10. Doi: 10.1186/1743-422X-6-10

19 Panning M, Grywna K, van Esbroeck M, Emmerich P, Drosten C. Chikungunya fever in travelers returning to Europe from the Indian Ocean region, 2006. Emerg Infect Dis 2008; 14(3): 416–422. doi: 10.3201/eid1403.070906

20 Lanciotti RS, Kosoy OL, Laven JJ, et al. Chikungunya virus in US travelers returning from India, 2006. Emerg Infect Dis 2007; 13(5): 764–767. doi: 10.3201/eid1305.070015

21 Stachler E, Kelty C, Sivaganesan M, Li X, Bibby K, Shanks OC. Quantitative crAssphage PCR assays for human fecal pollution measurement. Environ Sci Technol 2017; 51: 9146–9154.

22 Ahmed W, Payyappat S, Cassidy M, Besley C. A duplex PCR assay for the simultaneous quantification of Bacteroides HF183 and crassphage CPQ_056 marker genes in untreated sewage and stormwater. Environ Int 2019; 126: 252–259. doi: 10.1016/j.envint.2019.01.035

23 Cuevas-Ferrando E, Pérez-Cataluña A, Falcó I, Randazzo W, Sánchez G. Monitoring human viral pathogens reveals potential hazard for treated wastewater discharge or reuse. Front Microbiol 2022; 13: 836193. doi: 10.3389/fmicb.2022.836193

24 Irekeola AA, Syafirah EA, Islam MdA, Shueb RH. Global prevalence of dengue and chikungunya coinfection: a systematic review and meta-analysis of 43,341 participants. Acta Trop 2022; 231: 106408. doi: 10.1016/j.actatropica.2022.106408

25 Giunti G, Becker N, Benelli G. Invasive mosquito vectors in Europe: from bioecology to surveillance and management. Acta Trop 2023; 239: 106832. doi: 10.1016/j.actatropica.2023.106832

26 Soller J, Jennings W, Schoen M., et al. Modelling infection from SARS-CoV-2 wastewater concentrations: promise, limitations, and future directions. J Water Health 2022; 20: 1197–1211. Doi: 10.2166/wh.2022.094.

27 Huisman JS, Scire J, Caduff L, et al. Wastewater-base estimation of the effective reproductive number of SARS-CoV-2. Environ. Health Perspect 2022; 130: 57011. doi: 10.1289/EHP10050

